# Comparison of Mammography Artificial Intelligence Algorithms for 5-year Breast Cancer Risk Prediction

**DOI:** 10.1101/2022.01.05.22268746

**Authors:** Vignesh A. Arasu, Laurel A. Habel, Ninah S. Achacoso, Diana S.M. Buist, Jason B. Cord, Laura J. Esserman, Nola. M. Hylton, M. Maria Glymour, John Kornak, Lawrence H. Kushi, Don A. Lewis, Vincent X. Liu, Diana L. Miglioretti, Daniel A. Navarro, Weiva Sieh, Li Shen, Oleg Sofrygin, Hyo-Chun Yoon, Catherine Lee

## Abstract

**PURPOSE:** To examine the ability of 5 artificial intelligence (AI)-based computer vision algorithms, most trained to detect visible breast cancer on mammograms, to predict future risk relative to the Breast Cancer Surveillance Consortium clinical risk prediction model (BCSC v2).

**PATIENTS AND METHODS:** In this case-cohort study, women who had a screening mammogram in 2016 at Kaiser Permanente Northern California with no evidence of cancer on final imaging assessment were followed through September 2021. Women with prior breast cancer or a known highly penetrant gene mutation were excluded. From the 329,814 total eligible women, a random subcohort of 13,881 women (4.2%) were selected, of whom 197 had incident cancer. All 4,475 additional incident cancers were also included. Continuous AI-predicted scores were generated from the index 2016 mammogram. Risk estimates were generated with the Kaplan-Meier method and time-varying area under the curve [AUC(t)].

**RESULTS:** For incident cancers at 0-1 year (interval cancer risk), BCSC demonstrated an AUC(t) of 0.62 (95% CI, 0.58-0.66), and the AI algorithms had AUC(t)s ranging from 0.66-0.71, all significantly higher than BCSC (*P* < .05). For incident cancers at 1 to 5 years (5-year future cancer risk), BCSC demonstrated an AUC(t) of 0.61 (95% CI, 0.60-0.62), and the AI algorithms had AUC(t)s ranging from 0.63 to 0.67, all significantly higher than BCSC. Combined BCSC and AI models demonstrated AUC(t)s for interval cancer risk of 0.67-0.73 and for 5-year future cancer risk of 0.66-0.68.

**CONCLUSION:** The AI mammography algorithms we evaluated had significantly higher discrimination than the BCSC clinical risk model for interval and 5-year future cancer risk. Combined AI and BCSC models had slightly higher discrimination than AI alone.

## Introduction

Breast cancer risk prediction models are used to evaluate and guide clinical considerations such as hereditary risk, supplemental screening, and risk-reducing medications.^1^ Risk models are also under active investigation for broader population management, such as risk-based personalized screening^2, 3^ or capacity management.^4^ Several models have been developed to assess the risk for breast cancer in the general population, including Breast Cancer Risk Assessment Tool (BCRAT, also known as Gail),^5^ Breast Cancer Surveillance Consortium (BCSC),^6, 7^ and International Breast Cancer Intervention Study (also known as Tyrer-Cuzick).^8^ Beyond age, these models include clinical factors (e.g., family history of breast cancer, race/ethnicity, prior benign breast biopsy), genetic factors, and mammographic breast density. However, these models have only moderate discrimination for predicting either 5- or 10-year risk of breast cancer, with areas under the curve (AUC) ranging from 0.62 to 0.66.

Computer vision–based artificial intelligence (AI) models have the potential to improve risk prediction beyond clinical risk factors. These models quantitatively extract imaging biomarkers that represent underlying pathophysiologic mechanisms and phenotypes.^9^ Breast density is the single imaging biomarker most commonly incorporated into clinical risk models, but recent advances in AI deep learning^10^ provide the ability to extract hundreds to thousands of additional mammographic features beyond breast density alone. However, most mammography-based AI algorithms have been explicitly trained to assist a radiologist with detecting cancer visible on screening mammography (i.e., over a short time horizon) and not trained to help predict future risk several years after the time of examination.^11^ A few studies have evaluated future risk performance for AI algorithms explicitly trained for this task; these studies suggest substantial improvements over clinical risk models alone.^12, 13^ It is unknown whether AI trained for detection at shorter time horizons—the majority of mammography AI algorithms—can also predict longer-term risk.

We evaluated 5 commercial and academic mammography AI algorithms, trained for various time horizons, for their predictive performance at a 5-year time horizon following a negative mammogram in a large, community-based US cohort from the Kaiser Permanente Northern California (KPNC) integrated health system. We compared AI performance to the well-validated BCSC clinical risk model and explored whether combining the AI and BCSC risk models can further improve risk prediction above either model type alone.

## Methods

### Study Design, Setting, and Population

We performed a retrospective case-cohort study of women who had a bilateral screening mammogram in 2016 at KPNC (i.e., the index mammogram), without evidence of cancer on final imaging assessment either at the time of screening or after diagnostic work-up of positive screening findings. Women were excluded if they had a prior history of breast cancer or a high-penetrance breast cancer susceptibility gene as defined by the National Comprehensive Cancer Network guidelines.^14^ Of 329,814 women who met these criteria, a random subcohort of 13,881 women (4.2%) containing 197 cases was selected for analyses; all 4,475 additional incident cases diagnosed within 5 years of the index 2016 mammogram were also included (4,672 total; 100% of cases) (Figure 1). This sample size was based on the maximum cohort size feasible for AI algorithm evaluation. Our study was approved by the KPNC institutional review board for HIPAA compliance. The study followed the Strengthening the Reporting of Observational Studies in Epidemiology (STROBE) guidelines.^15, 16^

**Figure 1:**
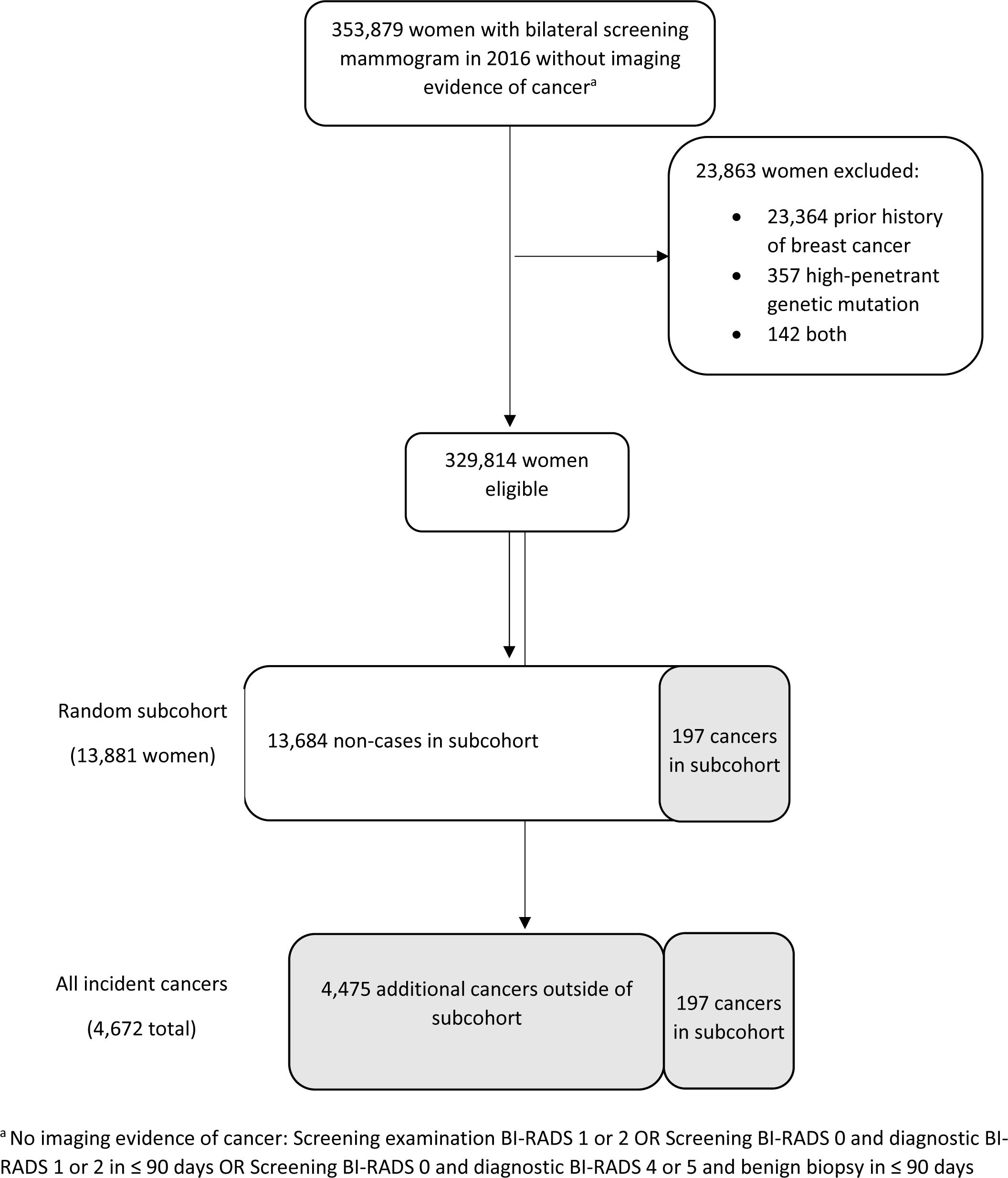
Case cohort selection

### Primary Outcome Ascertainment: Breast Cancer

The primary outcome was incident breast cancer defined as pathologically confirmed invasive carcinoma or ductal carcinoma in situ (DCIS). Cancers were ascertained from the KPNC Breast Cancer Tracking System^17^ quality assurance program, which has 99.8% concordance with the KPNC tumor registry that reports to the National Cancer Institute’s Surveillance, Epidemiology, and End Results (SEER) Program, but the Tracking System identifies incident cancers more rapidly (within 1 month of diagnosis) with manual verification. Women were followed from their index mammogram to date of breast cancer diagnosis; death; health plan disenrollment (allowing up to a 3-month gap in enrollment); or August 31, 2021, whichever occurred first.

### Primary Predictor Data Source: Negative Screening Mammograms from 2016

Screening mammograms in 2016 were identified by a Current Procedural Terminology examination code of 77057. A screening mammogram was considered to have no evidence of cancer on final assessment if the Breast Imaging Reporting and Data System (BI-RADS) assessment category was 1 or 2 on the screening mammogram, BI-RADS 0 on the screening mammogram and BI-RADS 1 or 2 on the diagnostic mammogram within 90 days, or BI-RADS 0 on the screening mammogram and BI-RADS 4 or 5 on the diagnostic mammogram with concordant benign biopsy within 90 days. The mammograms were evaluated in their archived processed form and were predominately (87%) acquired on Hologic mammography units (Hologic, Inc., Malborough MA), with the remainder acquired on General Electric units (GE Healthcare, Boston MA). In the KPNC health system, most average-risk women start screening mammography at age 50 years with a screening frequency of every 2 years, although women are given the option to screen starting at age 40 years or to screen annually.

### Primary Predictor Measurement: AI Risk Score Derived from Screening Mammogram

AI scores were generated from 5 deep-learning computer vision algorithms that take the screening mammogram images as their input, and output patient-level predicted scores. Candidate algorithms were chosen from an ongoing institutional AI operational evaluation. We evaluated 2 academic algorithms freely available for research, the Mirai algorithm (Massachusetts Institute of Technology, Boston, Massachusetts; https://www.github.com/yala/Mirai)^13^ and the Globally Aware Multiple Instance Classifier (GMIC) algorithm (New York University, New York, New York; https://www.github.com/nyukat/gmic). The 3 commercial vendor identities were anonymized for confidentiality and labeled Vendor A through Vendor C. While all but one of the algorithms were trained for substantially shorter time horizons, but we evaluated each algorithm’s ability to predict future risk up to 5 years. Further details of the software architecture for each algorithm are provided in the Data Supplement. Because the Mirai algorithm was also calibrated to provide predicted 5-years, further evaluation of calibration was performed for the Mirai algorithm and the BCSC risk score. When any algorithm failed to process an individual mammogram, this missing score was imputed using the algorithm’s specific overall median score (missingness by algorithm is detailed in Table S1 [Data Supplement]; evaluation by scored cases only is provided in Table S2).

### Comparative Predictor Ascertainment: Clinical Risk Score

The BCSC clinical 5-year risk prediction model version 2^6, 18^ was used as the comparator for the AI models. The BCSC model predicts risk for women without a history of breast cancer based on age, race/ethnicity, first-degree family history of breast cancer, prior benign breast biopsy, and mammographic breast density. To reflect the underlying population, clinical data for risk score generation were obtained in the KPNC electronic health record at or before the index date of the first screening mammogram in 2016, regardless of prior membership in the KPNC health system. Breast density was based on the index mammogram using the BI-RADS classification system by radiologists at the time of interpretation. Data completeness for family history and prior history of breast cancer depended on patient responses to clinic intake forms or notes by the provider during routine care; our data structure does not distinguish a woman with no family history from a woman with missing data. Breast biopsy data were available if obtained while the woman was enrolled in the KPNC health plan. Although our biopsy database prospectively classifies atypia and lobular carcinoma in situ, it does not distinguish proliferative benign pathology from otherwise benign pathology, so these benign outcomes were conservatively classified as non-proliferative lesions. Although the Mirai algorithm can input clinical variables for a combined risk score, at the time of evaluation this feature was not useable because it did not allow a variable number of missing risk factors.

### Statistical Analysis

R version 4.0.2^19^ was used for all statistical analyses. All statistical tests were 2-sided, with a threshold for statistical significance of α = .05. We evaluated the ability of the measures to predict breast cancer occurring within 3 time periods following mammography: “interval cancer risk” as incident cancers diagnosed between 0 and 1 years, “future cancer risk” as incident cancers diagnosed between 1 and 5 years, and “overall 5-year risk” as incident cancers diagnosed between 0 and 5 years.

Kaplan-Meier estimators were used to estimate the overall 5-year cumulative incidence of breast cancer within strata of each risk score (>90% percentile, middle 80%, <10% percentile); design weights were included to account for the case-cohort sampling. Discrimination was evaluated through the time-dependent AUC [AUC(t)], which accounts for the dynamic definition of cases and non-cases when handling time-to-event outcomes.^20^ We implemented the estimator that accounts for the censoring and sampling distribution using inverse probability of censoring weights and the case-cohort sampling^21^ and obtained corresponding 95% confidence intervals (CIs) using bootstrapping with 1000 bootstrap samples.^22^ To compare AUC(t) estimates from 2 separate risk scores, we calculated the difference in estimates and corresponding bootstrapped 95% CIs; a CI that does not contain 0 indicates that the difference in AUC(t) estimates is statistically significant at the .05 level.^23^

A Cox model was fit to predict 5-year risk by using the combined AI-predicted score and the BCSC score. The Cox models accounted for the case-cohort sampling through design weights and included both the AI score and BCSC score flexibly using restricted cubic splines with 4 knots.^24, 25^ We used 5-fold cross-validation (CV) to estimate the AUC(t) estimator described above^21^ and present the average value across the 5-folds. We obtained corresponding 95% CIs for the average CV-AUC(t) through bootstrapping with 1000 bootstrap samples.

We assessed the 5-year calibration of Mirai and BCSC risk within prespecified strata of 5-year risk (0 to <1.67%, 1.67 to <3%, ≥3%) based on thresholds established by the BCSC. We compared the observed number of cases over the 5-year study period with the expected number of cases, calculated as the sum of the cumulative hazard estimates over all individuals in the study.^26^ We report the ratio of observed to expected cases with exact 95% CIs.^27^ We calculated the incidence rates (IRs; cases per 1000 person-years) and IR ratios (IRRs) with 95% CIs based on a Poisson distribution.^28^ All expected incidence estimates incorporated design weights that account for the case-cohort sampling.

## Results

### Patient Characteristics

Although our population was predominately older, non-Hispanic white women, a substantial proportion of the women were younger than age 50 years (23%) or of non-White race/ethnicity (48%) (Table 1). Women had a median of 5.0 years of follow-up (interquartile range 4.7 to 5.3 years) and were censored due to end of follow-up (92%), disenrollment (6%), or death (2%).

**Table 1:** Patient characteristics

|  | Women in<br>subcohort, n (%) | Women with<br>breast cancer, n<br>(%)* | All eligible women,<br>n (%) |
| --- | --- | --- | --- |
| Total women | 13 881 (100) | 4672 (100) | 329 814 (100) |
| Age, years |  |  |  |
| <40 | 84 (1) | 19 (<1) | 2011 (1) |
| 40-49 | 3149 (23) | 735 (16) | 74 887 (21) |
| 50-59 | 4792 (35) | 1331 (28) | 114 780 (33) |
| 60-69 | 4215 (30) | 1809 (39) | 99 341 (32) |
| ≥70 | 1641 (12) | 778 (17) | 38 795 (13) |
| Race/Ethnicity |  |  |  |
| Black, non-Hispanic | 976 (7) | 329 (7) | 23 721 (7) |
| Asian or Pacific Islander | 2672 (19) | 897 (19) | 63 228 (19) |
| Hispanic | 2401 (17) | 572 (12) | 57 389 (17) |
| Multiracial | 504 (4) | 163 (3) | 11 079 (3) |
| Native American | 53 (<1) | 18 (<1) | 1392 (<1) |
| White, non-Hispanic | 7037 (51) | 2676 (57) | 167 781 (51) |
| Missing | 238 (2) | 17 (<1) | 5224 (2) |
| First-degree family history |  |  |  |
| 0 | 12 150 (88) | 3750 (80) | 288 317 (87) |
| 1 | 1639 (12) | 866 (19) | 39 463 (12) |
| ≥ 2 | 92 (1) | 56 (1) | 2034 (1) |
| Previous benign breast biopsies |  |  |  |
| 0 | 13 119 (95) | 4160 (89) | 310 805 (94) |
| ≥1 | 762 (5) | 512 (11) | 19009 (6) |
| BI-RADS breast density |  |  |  |
| Almost entirely fat | 1405 (10) | 250 (5) | 30 886 (9) |
| Scattered fibroglandular<br>densities | 6387 (46) | 2014 (43) | 153 752 (47) |
| Heterogeneously dense | 5341 (38) | 2117 (45) | 125 855 (38) |
| Extremely dense | 748 (5) | 251 (5) | 17 596 (5) |
| Missing | 65 (<1) | 40 (1) | 1725 (1) |
| Cancer type |  |  |  |
| Invasive | 152 (77) | 3850 (82) | 3850 (82) |
| DCIS | 45 (23) | 822 (18) | 822 (18) |
| Median follow-up interval, years<br>(interquartile range) | 5.0 (4.7 to 5.3) | 2.8 (2.0 to 4.1) | 5.0 (4.7 to 5.3) |
| Median healthcare enrollment prior<br>to index date, years (interquartile<br>range) | 17.9 (9.7 to 19.4) | 18.9 (10.7 to 19.5) | 17.6 (9.2 to 19.4) |

\*Includes cases within subcohort (n=197) as well as cases outside subcohort (n=4,475)

### Cumulative Incidence Rates of BCSC Clinical Risk Model and AI Algorithm Scores

The BCSC average cumulative IR at 5 years in women with risk score percentile >90% was 30.3 per 1000 (95% CI, 28.0-32.9 per 1000), middle 80% was 15.0 per 1000 (95% CI, 14.5-15.7 per 1000), and <10% was 6.1 per 1000 (95% CI, 5.1-7.2 per 1000) (Figure 2). The IRR of >90% risk to <10% risk was 5.4. Women with a BCSC risk >90% predicted 21% of all cancers by 5 years, whereas women < 10% predicted 3% of all cancers.

**Figure 2:**
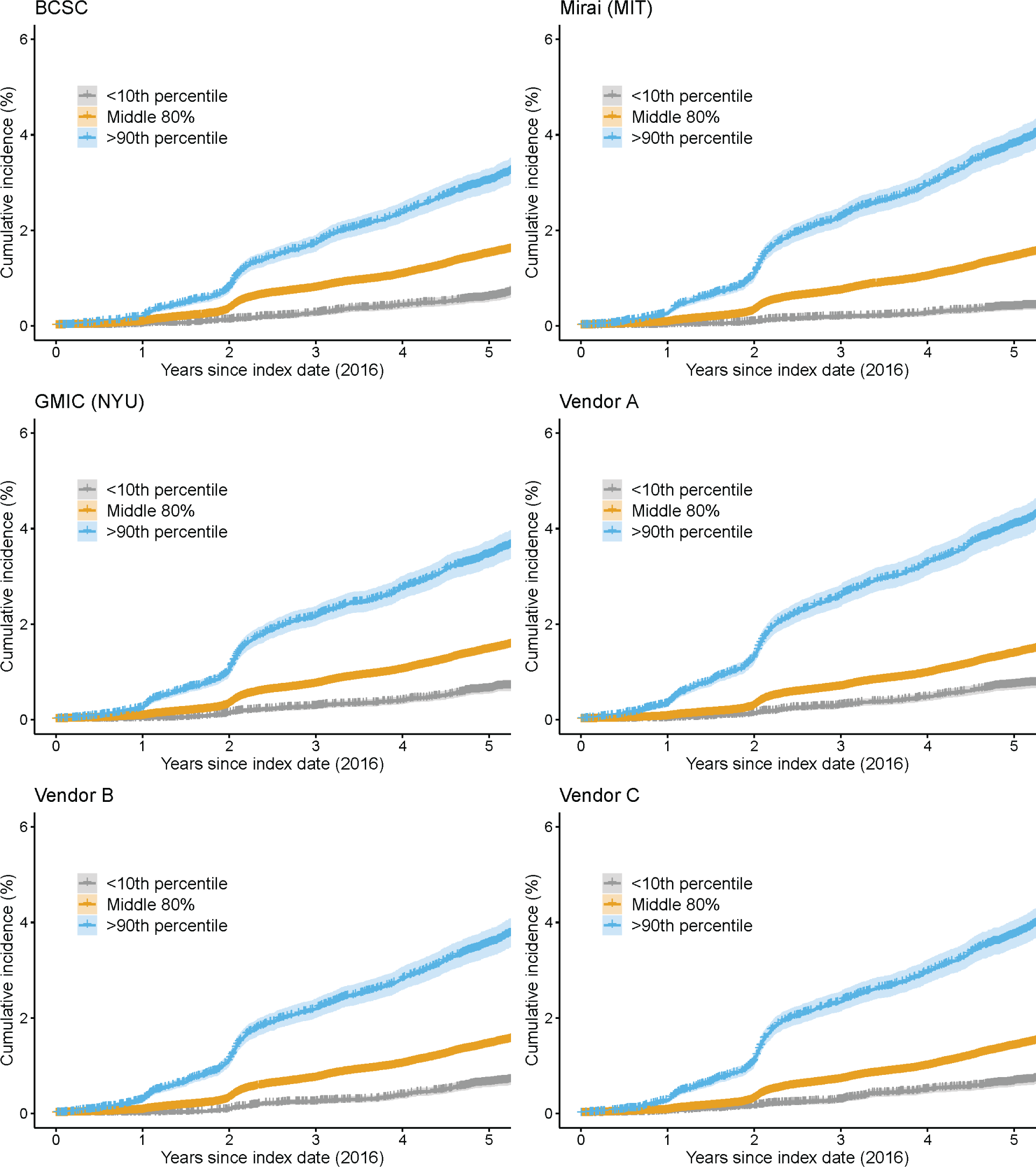
Cumulative risk of breast cancer by AI score at 5-years

For AI algorithms, the average cumulative IR at 5 years in women with risk score percentile >90% ranged from 31.3 to 40.9 per 1000, middle 80%, from 13.7 to 15.0 per 1000, and <10% from 6.4 to 7.4 per 1000. The IRR of the >90% risk to <10% risk ranged between 5.3 and 7.3. Women with >90% risk predicted 20% to 27% of all cancers by 5 years, whereas women with <10% risk predicted approximately 2% to 4% of cancers across all AI algorithms.

### Discrimination and Calibration of BCSC Clinical Risk Model and AI Algorithm Scores

When evaluating discrimination for interval cancer risk (Table 2), BCSC demonstrated an AUC(t) of 0.62 (95% CI, 0.58-0.66), whereas the AI algorithms AUC(t)s ranged between 0.66 to 0.71, all significantly higher than BCSC (*P* < .05). For 5-year future cancer risk, BCSC demonstrated an AUC(t) of 0.61 (95% CI, 0.60-0.62), whereas the AI algorithm AUC(t)s ranged between 0.61 to 0.67, with all algorithms significantly higher than BCSC (*P* < .05).

**Table 2:**
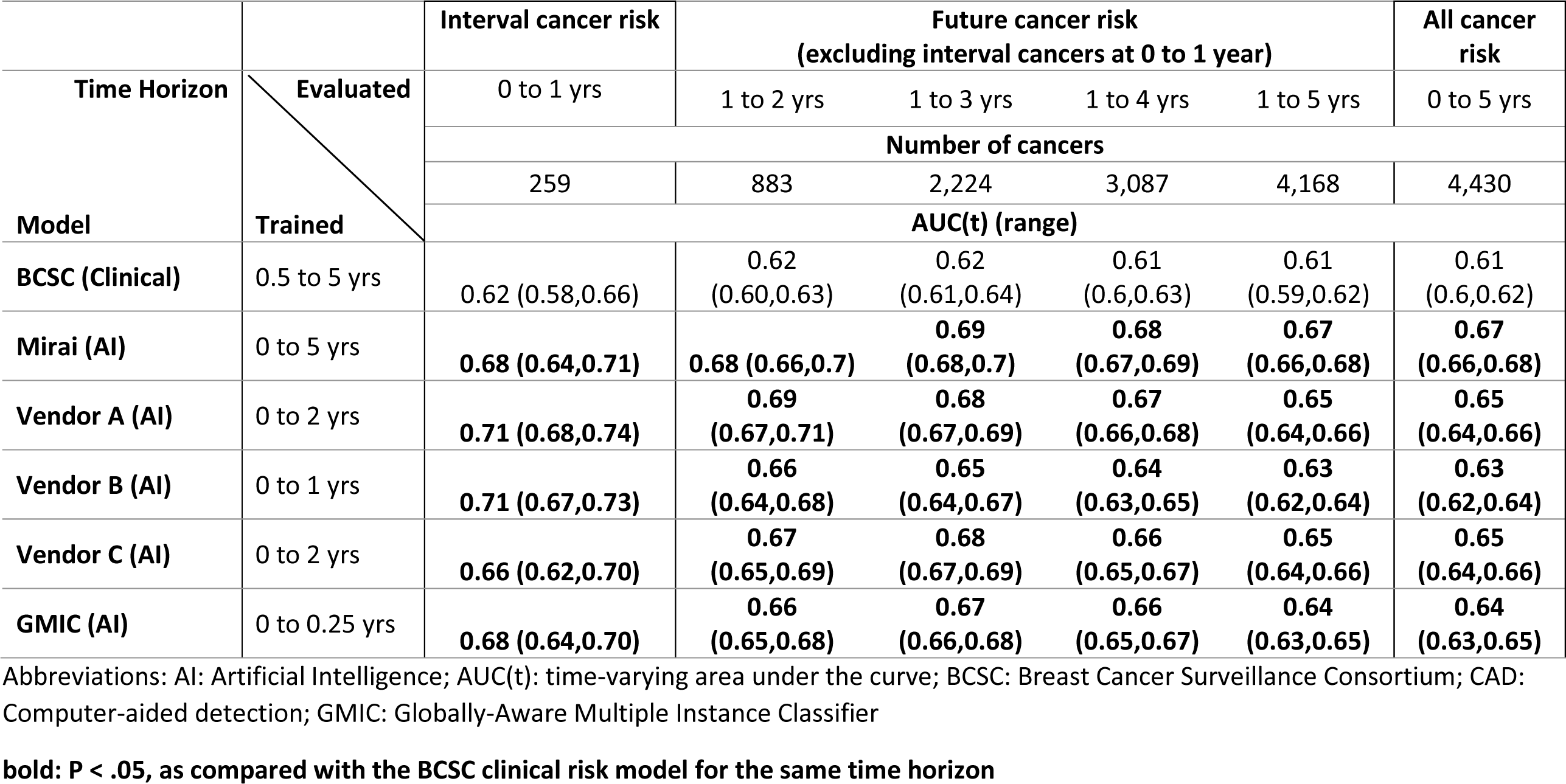
Comparative AUC(t) performance of AI models and BCSC clinical risk model using 2016 screening mammogram without evidence of cancer on final imaging assessment for invasive cancer and DCIS.

| Time Horizon | Evaluated | Interval cancer risk | Future cancer risk<br>(excluding interval cancers at 0 to 1 year) |  |  |  | All cancer risk |
| --- | --- | --- | --- | --- | --- | --- | --- |
|  |  | 0 to 1 yrs | 1 to 2 yrs | 1 to 3 yrs | 1 to 4 yrs | 1 to 5 yrs | 0 to 5 yrs |
| Model | Trained | Number of cancers |  |  |  |  |  |
|  |  | 259 | 883 | 2,224 | 3,087 | 4,168 | 4,430 |
|  |  | AUC(t) (range) |  |  |  |  |  |
| BCSC (Clinical) | 0.5 to 5 yrs | 0.62 (0.58,0.66) | 0.62 (0.60,0.63) | 0.62 (0.61,0.64) | 0.61 (0.6,0.63) | 0.61 (0.59,0.62) | 0.61 (0.6,0.62) |
| Mirai (AI) | 0 to 5 yrs | **0.68 (0.64,0.71)** | **0.68 (0.66,0.7)** | **0.69 (0.68,0.7)** | **0.68 (0.67,0.69)** | **0.67 (0.66,0.68)** | **0.67 (0.66,0.68)** |
| Vendor A (AI) | 0 to 2 yrs | **0.71 (0.68,0.74)** | **0.69 (0.67,0.71)** | **0.68 (0.67,0.69)** | **0.67 (0.66,0.68)** | **0.65 (0.64,0.66)** | **0.65 (0.64,0.66)** |
| Vendor B (AI) | 0 to 1 yrs | **0.71 (0.67,0.73)** | **0.66 (0.64,0.68)** | **0.65 (0.64,0.67)** | **0.64 (0.63,0.65)** | **0.63 (0.62,0.64)** | **0.63 (0.62,0.64)** |
| Vendor C (AI) | 0 to 2 yrs | **0.66 (0.62,0.70)** | **0.67 (0.65,0.69)** | **0.68 (0.67,0.69)** | **0.66 (0.65,0.67)** | **0.65 (0.64,0.66)** | **0.65 (0.64,0.66)** |
| GMIC (AI) | 0 to 0.25 yrs | **0.68 (0.64,0.70)** | **0.66 (0.65,0.68)** | **0.67 (0.66,0.68)** | **0.66 (0.65,0.67)** | **0.64 (0.63,0.65)** | **0.64 (0.63,0.65)** |

Abbreviations: AI: Artificial Intelligence; AUC(t): time-varying area under the curve; BCSC: Breast Cancer Surveillance Consortium; CAD: Computer-aided detection; GMIC: Globally-Aware Multiple Instance Classifier
**bold: P < .05, as compared with the BCSC clinical risk model for the same time horizon**

AI and BCSC models were combined using restricted cubic splines due to violation of proportional hazards assumption as linear models. Combined model AUC(t)s for interval cancer risk ranged between 0.67 to 0.73 (Table 3) and were only significantly higher than the corresponding AI algorithm alone for Mirai and GMIC. The combined model AUC(t) for 5-year future cancer risk ranged from 0.65 to 0.68 and were significantly higher for all combined models.

**Table 3:**
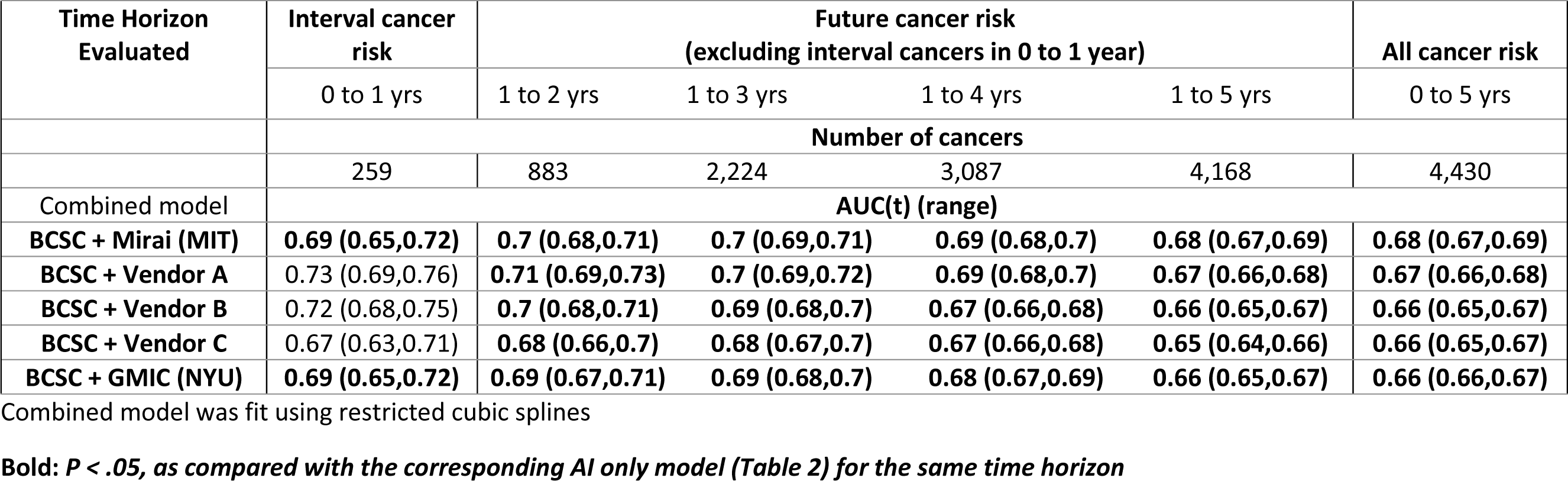
Comparative time-varying AUC(t) performance of combined AI models and BCSC clinical risk model using 2016 screening mammogram without evidence of cancer on final imaging assessment.

Additional subgroup analyses (Tables S2-S7, Data Supplement) demonstrated comparable discrimination to that shown in Table 2 when restricted to women with invasive breast cancer, with complete scores available across all models, with BI-RADS 1 or 2 on screening mammograms, and with mammograms acquired on Hologic equipment. Discrimination was mixed for women with BI-RADS 0 on screening mammograms or mammograms acquired on GE equipment only.

The 5-year calibration of the BCSC ranged from 1.02 to 1.07 depending on the prespecified risk threshold ranges, whereas that of the Mirai algorithm ranged from 0.49 to 0.76 (Table 4).

**Table 4:** Absolute risk calibration by Mirai AI and BCSC clinical risk models at 5-year cumulative incidence thresholds

|  |  |  | No. of Cancer Cases |  |  | IR per 1000 women/y |  |  |
| --- | --- | --- | --- | --- | --- | --- | --- | --- |
| Model by 5-year Risk | No. (%) of women* | Follow-up, 1000 women years (weighted) | Observed | Expected (weighted) | O/E (95% CI) | Observed | Expected (weighted) | Observed IRR (95% CI) |
| BCSC |  |  |  |  |  |  |  |  |
| All | 18 356 | 1422.7 | 4430 | 4172.4 | 1.06 (1.03,1.09) | 3.11 | 2.93 |  |
| 0- <1.67% | 14 499 | 1164.5 | 3048 | 2825.2 | 1.08 (1.04,1.12) | 2.62 | 2.43 | 1.00 |
| 1.67- <3% | 3387 | 233.3 | 1150 | 1129.5 | 1.02 (0.96,1.08) | 4.93 | 4.84 | 1.88 (1.76,2.02) |
| ≥3% | 470 | 25.0 | 232 | 217.7 | 1.07 (0.93,1.21) | 9.28 | 8.71 | 3.55 (3.10,4.05) |
| Mirai |  |  |  |  |  |  |  |  |
| All | 18 356 | 1422.7 | 4430 | 7360.5 | 0.60 (0.58,0.62) | 3.11 | 5.17 |  |
| 0- <1.67% | 7597 | 672.4 | 1010 | 2060.1 | 0.49 (0.46,0.52) | 1.50 | 3.06 | 1.00 |
| 1.67- <3% | 8042 | 592.6 | 2233 | 2935.9 | 0.76 (0.73,0.79) | 3.77 | 4.95 | 2.51 (2.33,2.70) |
| ≥3% | 2717 | 157.6 | 1187 | 2364.5 | 0.5 (0.47,0.53) | 7.53 | 15.00 | 5.01 (4.61,5.45) |

Abbreviations: O/E: Observed to expected ratio; IR: Incidence rate; IRR: Incidence rate ratio for observed IRs; BCSC: Breast Cancer Surveillance Consortium
\*Includes 13,881 women in subcohort plus 4,475 additional women with cancer

## Discussion

In this comparative assessment of breast cancer risk models, all AI algorithms had significantly higher discrimination than the BCSC clinical risk model for predicting 5-year risk. This difference was most pronounced for interval cancer risk. Furthermore, we demonstrated that AI algorithms mostly trained for short time horizons can predict future risk of cancer up to 5 years when no cancer is clinically detected on mammography. Combining BCSC and AI further improves risk prediction above AI alone, and decreases the gap in future risk performance between AI algorithms.

Mammography AI algorithms provide a new approach for improving breast cancer risk prediction beyond clinical variables such as age, family history, or the traditional imaging risk biomarker of breast density. The absolute increase in the AUC for the best mammography AI relative to BCSC was 0.09 for interval cancer risk and 0.06 for overall 5-year risk, a substantial and clinically meaningful improvement. This improvement also remained when restricting to invasive cancer only. Improved interval cancer risk prediction is expected, given that most AI are trained for short time horizons to aid radiologists with detecting cancers. However, continued strong predictive performance up to 5 years is surprising and suggests AI is no longer identifying missed cancers, but features of true underlying risk. This is analogous to breast density predicting interval cancer risk due to tissue masking cancers, but also predicting future risk where masking is no longer contributory^30^.

The combined AI and clinical risk model marginally improved performance compared with any AI model alone. This incremental improvement was also noted in other studies combining mammography AI and clinical risk.^12, 13^ The combined model also decreased overall differences in discrimination between AI algorithms. Larger gains in improvement may be derived by combining clinical risk and mammography AI with single nucleotide polymorphism polygenic risk scores,^12^ which we will evaluate in the future.

To understand pragmatic performance at population level, we used a relatively unselected cohort and incorporated median imputation when AI failed to generate a risk score (although most algorithms had a very low failure rate). We also evaluated risk at different time horizons because each has distinct clinical implications. AI algorithms particularly excel at predicting high risk of interval cancer, which is associated with aggressive cancers^30, 31^ and may lead to second reading of mammograms, supplementary screening (e.g., with breast MRI), or short-interval follow-up. AI algorithms also predict elevated future risk, which may lead to more frequent and intensive screening or risk counseling for primary prevention.

The BCSC model prediction was originally calibrated to US SEER incidence rates and remained well calibrated in our cohort, confirming that our population is likely representative of community-based populations. In contrast, the Mirai model overestimated cancer risk by a factor of 2 across all risk strata. The Mirai predicted risk was originally calibrated using women from a tertiary referral setting who likely had a higher cancer rate than the women in our health system. Although calibration does not affect the observed discriminative performance, it is critical when clinical decisions are based on prespecified risk model thresholds. However, given its systematic overestimation, the Mirai model may be recalibrated for these purposes.

Beyond improved performance, mammography-based AI risk models provide practical advantages over traditional clinical risk models. AI uses a single data source (the screening mammogram) that is available for most women for whom breast cancer risk prediction is relevant, enabling risk scores to be generated consistently and efficiently for all women in a large population. Mammography AI risk models overcome certain barriers for risk models such as time and cost for combining multiple data elements from potentially different sources, as well as dependence on patient-reported history and susceptibility to missing data or recall bias. However, mammography AI risk models also have potential costs (e.g., for new software or graphics processing hardware) and other new technical and workflow considerations for implementation. Some breast imaging practices may already incorporate mammography AI trained for aiding immediate detection, and the generated score can simultaneously be used for future risk stratification. However, before AI is applied, it should be evaluated in specific populations that are likely to experience health disparities if there are hidden biases in the algorithms.^32^

We evaluated our results in a diverse, community-based cohort, using a rigorous design and methods to evaluate AI under both pragmatic and optimal conditions. Our observed discrimination was consistent with prior publications for the Mirai AI algorithm^13^ and the BCSC clinical risk model.^6, 7, 33^ Our results are not an endorsement of any one AI algorithm, but a demonstration of the inherent predictive power in mammography-based deep learning using a sample of 5 AI algorithms. It is beyond the scope of our study to evaluate the hundreds of mammography AI algorithms available at this time,^11^ but similar results may be seen in algorithms not evaluated in our study.

Our study was limited due to retrospective ascertainment of BCSC clinical risk model inputs for family history and prior breast biopsies. We are unable to assess the extent to which these data are missing, particularly for breast biopsies performed prior to enrollment in our health plan. Although prevalence of family history was comparable to BCSC estimates, a history of breast biopsy was 10-15% lower than previously reported,^7^ which may have led to underestimation of BCSC performance.

Our results imply that mammography AI algorithms alone may be a clinically meaningful improvement over current risk models at early time horizons, with further improvements in prediction when AI and clinical risk models are combined. Although AI algorithm performance declines with longer time horizons, most of the algorithms evaluated have not yet been trained to predict longer-term outcomes, suggesting a rich opportunity for further improvement. Moreover, AI provides a powerful way to stratify women for clinical considerations that necessitate shorter time horizons, such as risk-based screening and supplemental imaging. The impact on clinical decisions requiring longer-term data, such as for chemoprevention or hereditary genetic screening, requires further study in cohorts with longer follow- up.

## Data Availability

All data produced in the present study are available upon reasonable request to the authors

## Acknowledgement

This research was supported by The Permanente Medical Group (TPMG) Delivery Science and Applied Research Physician Researcher Program. The authors thank Jane Bethard-Tracy and the Kaiser Permanente Northern California Breast Cancer Tracking System staff, Albert Pu, Bing Lee, Naomi Ruff, Caitlin Lydon, and Seth Selkow for their additional contributions in supporting this study. The authors also thank the participating women, mammography facilities, and radiologists for the data they provided for this study.

---

**Table S1.** supplement only: Missing data by model.

|  | **All observations** | **All missing (%)** | **Cases missing (%)** | **Non cases missing (%)** |
| --- | --- | --- | --- | --- |
| **BCSC** |  |  |  |  |
| **Mirai (MIT)** | 18356 | 602 (3.3) | 231 (4.9) | 371 (2.7) |
| **Vendor A** | 18356 | 479 (2.6) | 153 (3.3) | 326 (2.4) |
| **Vendor B** | 18356 | 2407 (13.1) | 679 (14.5) | 1728 (12.6) |
| **Vendor C** | 18356 | 625 (3.4) | 229 (4.9) | 396 (2.9) |
| **GMIC (NYU)** | 18356 | 352 (1.9) | 245 (5.2) | 107 (0.8) |

Abbreviations: AI: Artificial Intelligence; AUC(t): time-varying area under the curve; BCSC: Breast Cancer Surveillance Consortium; CAD: Computer-aided detection; GMIC: Globally-Aware Multiple Instance Classifier

**Table S2.** supplement only: Complete scored women across all models only.

|  | Cancer risk type | Interval cancer risk | Future cancer risk<br>(excluding interval cancers at 0 to 1 year) |  |  |  | All cancer risk |
| --- | --- | --- | --- | --- | --- | --- | --- |
| Time horizon | Evaluated (y) | 0 to 1 | 1 to 2 | 1 to 3 | 1 to 4 | 1 to 5 | 0 to 5 |
| Model | Trained (y) | AUC(t) (range) |  |  |  |  |  |
| **BCSC** | 0.5 to 5 yrs | 0.63<br>(0.58,0.66) | 0.62<br>(0.6,0.63) | 0.63<br>(0.61,0.64) | 0.62<br>(0.6,0.63) | 0.61<br>(0.6,0.62) | 0.61<br>(0.6,0.62) |
| **Mirai (MIT)** | 0 to 5 yrs | 0.69<br>(0.64,0.71) | 0.69<br>(0.66,0.7) | 0.69<br>(0.68,0.7) | 0.69<br>(0.67,0.69) | 0.68<br>(0.66,0.68) | 0.68<br>(0.66,0.68) |
| **Vendor A** | 0 to 2 yrs | 0.72<br>(0.68,0.75) | 0.69<br>(0.67,0.71) | 0.69<br>(0.67,0.69) | 0.67<br>(0.66,0.68) | 0.65<br>(0.64,0.66) | 0.66<br>(0.64,0.66) |
| **Vendor B** | 0 to 1 yrs | 0.73<br>(0.67,0.74) | 0.68<br>(0.64,0.68) | 0.67<br>(0.64,0.67) | 0.66<br>(0.63,0.65) | 0.64<br>(0.62,0.64) | 0.64<br>(0.62,0.64) |
| **Vendor C** | 0 to 2 yrs | 0.67<br>(0.63,0.7) | 0.68<br>(0.65,0.69) | 0.68<br>(0.67,0.69) | 0.67<br>(0.65,0.67) | 0.65<br>(0.64,0.66) | 0.65<br>(0.64,0.66) |
| **GMIC (NYU)** | 0 to 0.25 yrs | 0.68<br>(0.64,0.71) | 0.67<br>(0.64,0.68) | 0.67<br>(0.66,0.68) | 0.66<br>(0.65,0.67) | 0.64<br>(0.63,0.65) | 0.65<br>(0.63,0.65) |

Abbreviations: AI: Artificial Intelligence; AUC(t): time-varying area under the curve; BCSC: Breast Cancer Surveillance Consortium; CAD: Computer-aided detection; GMIC: Globally-Aware Multiple Instance Classifier

**Table S3.**
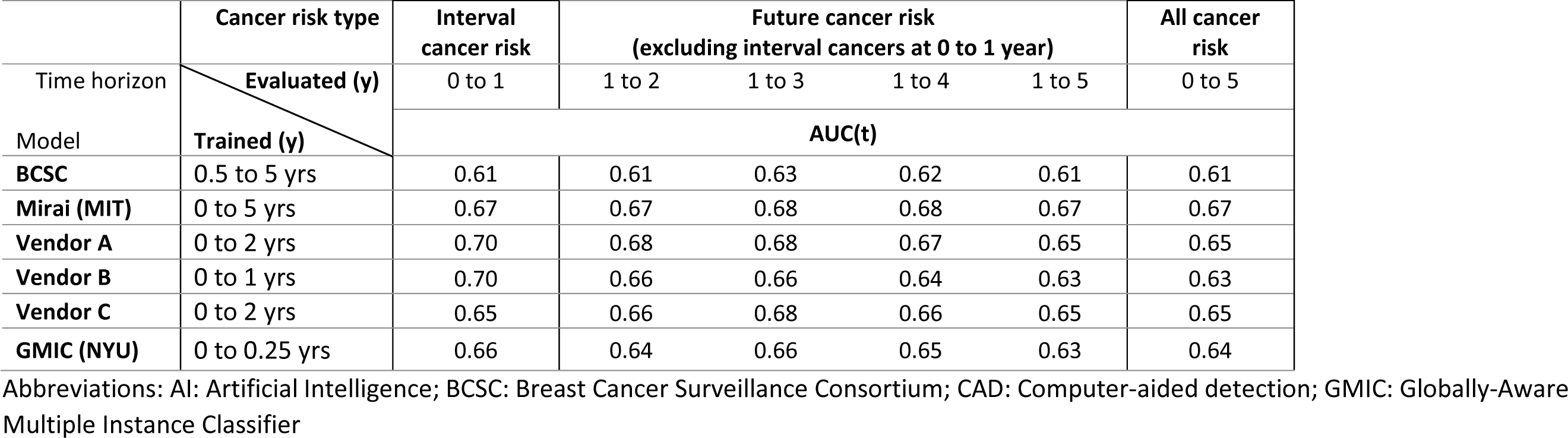
supplement only: Invasive cancer only. Comparative AUC(t) performance of AI models and BCSC clinical risk model for invasive cancer only using 2016 screening mammogram without evidence of cancer on final imaging assessment

**Table S4.** supplement only: Screening mammograms acquired on Hologic equipment, Complete cases only.

|  | Cancer risk type | Interval cancer risk | Future cancer risk<br>(excluding interval cancers at 0 to 1 year) |  |  |  | All cancer risk |
| --- | --- | --- | --- | --- | --- | --- | --- |
| Time horizon | Evaluated (y) | 0 to 1 | 1 to 2 | 1 to 3 | 1 to 4 | 1 to 5 | 0 to 5 |
| Model | Trained (y) | AUC(t) (range) |  |  |  |  |  |
| **BCSC** | 0.5 to 5 yrs | 0.63<br>(0.59,0.67) | 0.63<br>(0.6,0.65) | 0.63<br>(0.62,0.65) | 0.62<br>(0.61,0.63) | 0.61<br>(0.6,0.62) | 0.61<br>(0.6,0.62) |
| **Mirai (MIT)** | 0 to 5 yrs | 0.69<br>(0.65,0.72) | 0.69<br>(0.67,0.71) | 0.69<br>(0.68,0.71) | 0.69<br>(0.68,0.7) | 0.67<br>(0.66,0.69) | 0.68<br>(0.66,0.69) |
| **Vendor A** | 0 to 2 yrs | 0.72<br>(0.68,0.76) | 0.69<br>(0.67,0.72) | 0.68<br>(0.67,0.7) | 0.67<br>(0.66,0.68) | 0.65<br>(0.64,0.66) | 0.66<br>(0.65,0.67) |
| **Vendor B** | 0 to 1 yrs | 0.73<br>(0.69,0.77) | 0.68<br>(0.66,0.7) | 0.67<br>(0.66,0.69) | 0.66<br>(0.65,0.67) | 0.65<br>(0.63,0.66) | 0.65<br>(0.64,0.66) |
| **Vendor C** | 0 to 2 yrs | 0.67<br>(0.63,0.71) | 0.68<br>(0.65,0.7) | 0.68<br>(0.67,0.69) | 0.66<br>(0.65,0.68) | 0.65<br>(0.64,0.66) | 0.65<br>(0.64,0.66) |
| **GMIC (NYU)** | 0 to 0.25 yrs | 0.69<br>(0.65,0.72) | 0.67<br>(0.65,0.69) | 0.68<br>(0.66,0.69) | 0.67<br>(0.66,0.68) | 0.65<br>(0.64,0.66) | 0.66<br>(0.64,0.67) |

Abbreviations: AI: Artificial Intelligence; AUC(t): time-varying area under the curve; BCSC: Breast Cancer Surveillance Consortium; CAD: Computer-aided detection; GMIC: Globally-Aware Multiple Instance Classifier

**Table S5.**
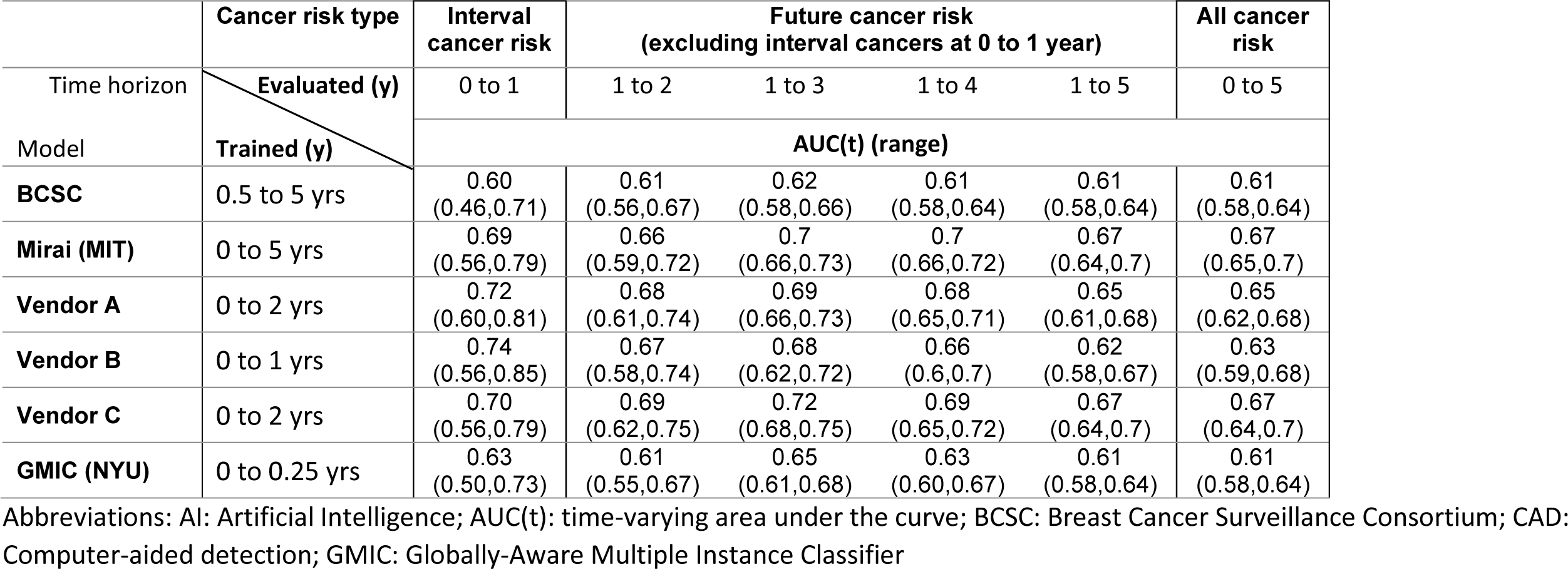
supplement only: Screening mammograms acquired on General Electric equipment, Complete cases only.

|  | Cancer risk type | Interval cancer risk | Future cancer risk<br>(excluding interval cancers at 0 to 1 year) |  |  |  | All cancer risk |
| --- | --- | --- | --- | --- | --- | --- | --- |
| Time horizon | Evaluated (y)<br>Trained (y) | 0 to 1 | 1 to 2 | 1 to 3 | 1 to 4 | 1 to 5 | 0 to 5 |
| Model |  | AUC(t) (range) |  |  |  |  |  |
| BCSC | 0.5 to 5 yrs | 0.60<br>(0.46,0.71) | 0.61<br>(0.56,0.67) | 0.62<br>(0.58,0.66) | 0.61<br>(0.58,0.64) | 0.61<br>(0.58,0.64) | 0.61<br>(0.58,0.64) |
| Mirai (MIT) | 0 to 5 yrs | 0.69<br>(0.56,0.79) | 0.66<br>(0.59,0.72) | 0.7<br>(0.66,0.73) | 0.7<br>(0.66,0.72) | 0.67<br>(0.64,0.7) | 0.67<br>(0.65,0.7) |
| Vendor A | 0 to 2 yrs | 0.72<br>(0.60,0.81) | 0.68<br>(0.61,0.74) | 0.69<br>(0.66,0.73) | 0.68<br>(0.65,0.71) | 0.65<br>(0.61,0.68) | 0.65<br>(0.62,0.68) |
| Vendor B | 0 to 1 yrs | 0.74<br>(0.56,0.85) | 0.67<br>(0.58,0.74) | 0.68<br>(0.62,0.72) | 0.66<br>(0.6,0.7) | 0.62<br>(0.58,0.67) | 0.63<br>(0.59,0.68) |
| Vendor C | 0 to 2 yrs | 0.70<br>(0.56,0.79) | 0.69<br>(0.62,0.75) | 0.72<br>(0.68,0.75) | 0.69<br>(0.65,0.72) | 0.67<br>(0.64,0.7) | 0.67<br>(0.64,0.7) |
| GMIC (NYU) | 0 to 0.25 yrs | 0.63<br>(0.50,0.73) | 0.61<br>(0.55,0.67) | 0.65<br>(0.61,0.68) | 0.63<br>(0.60,0.67) | 0.61<br>(0.58,0.64) | 0.61<br>(0.58,0.64) |

Abbreviations: AI: Artificial Intelligence; AUC(t): time-varying area under the curve; BCSC: Breast Cancer Surveillance Consortium; CAD: Computer-aided detection; GMIC: Globally-Aware Multiple Instance Classifier

**Table S6.**
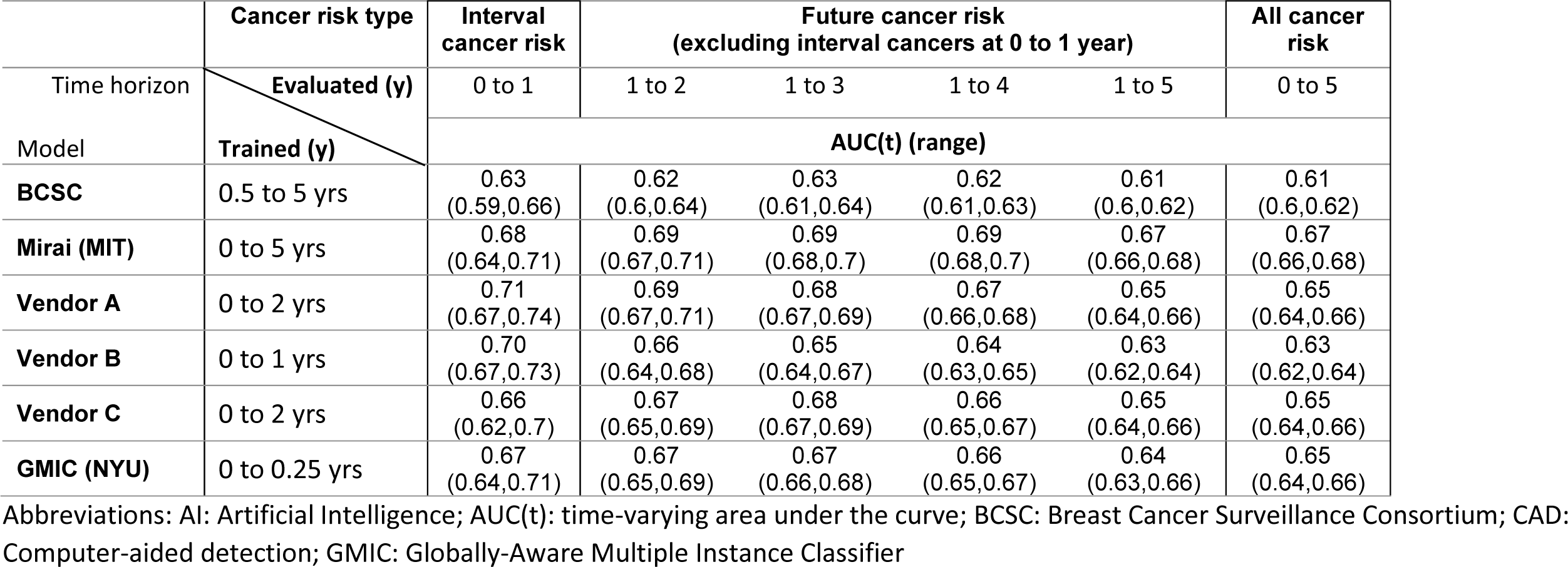
supplement only: Screening BI-RADS 1 or 2 only.

|  | Cancer risk type | Interval cancer risk | Future cancer risk<br>(excluding interval cancers at 0 to 1 year) |  |  |  | All cancer risk |
| --- | --- | --- | --- | --- | --- | --- | --- |
| Time horizon | Evaluated (y) | 0 to 1 | 1 to 2 | 1 to 3 | 1 to 4 | 1 to 5 | 0 to 5 |
| Model | Trained (y) | AUC(t) (range) |  |  |  |  |  |
| **BCSC** | 0.5 to 5 yrs | 0.63<br>(0.59,0.66) | 0.62<br>(0.6,0.64) | 0.63<br>(0.61,0.64) | 0.62<br>(0.61,0.63) | 0.61<br>(0.6,0.62) | 0.61<br>(0.6,0.62) |
| **Mirai (MIT)** | 0 to 5 yrs | 0.68<br>(0.64,0.71) | 0.69<br>(0.67,0.71) | 0.69<br>(0.68,0.7) | 0.69<br>(0.68,0.7) | 0.67<br>(0.66,0.68) | 0.67<br>(0.66,0.68) |
| **Vendor A** | 0 to 2 yrs | 0.71<br>(0.67,0.74) | 0.69<br>(0.67,0.71) | 0.68<br>(0.67,0.69) | 0.67<br>(0.66,0.68) | 0.65<br>(0.64,0.66) | 0.65<br>(0.64,0.66) |
| **Vendor B** | 0 to 1 yrs | 0.70<br>(0.67,0.73) | 0.66<br>(0.64,0.68) | 0.65<br>(0.64,0.67) | 0.64<br>(0.63,0.65) | 0.63<br>(0.62,0.64) | 0.63<br>(0.62,0.64) |
| **Vendor C** | 0 to 2 yrs | 0.66<br>(0.62,0.7) | 0.67<br>(0.65,0.69) | 0.68<br>(0.67,0.69) | 0.66<br>(0.65,0.67) | 0.65<br>(0.64,0.66) | 0.65<br>(0.64,0.66) |
| **GMIC (NYU)** | 0 to 0.25 yrs | 0.67<br>(0.64,0.71) | 0.67<br>(0.65,0.69) | 0.67<br>(0.66,0.68) | 0.66<br>(0.65,0.67) | 0.64<br>(0.63,0.66) | 0.65<br>(0.64,0.66) |

Abbreviations: AI: Artificial Intelligence; AUC(t): time-varying area under the curve; BCSC: Breast Cancer Surveillance Consortium; CAD: Computer-aided detection; GMIC: Globally-Aware Multiple Instance Classifier

**Table S7.**
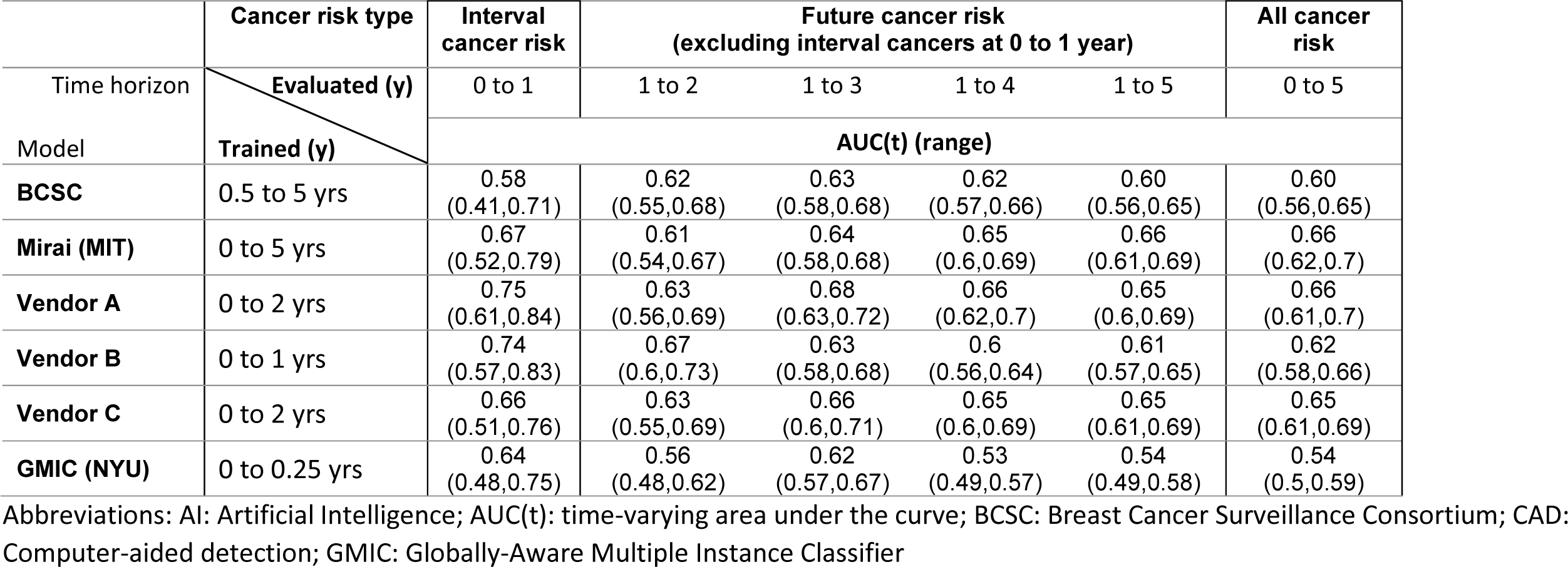
supplement only: Screening BI-RADS 0, and Diagnostic or post-procedural BI-RADS 1 or 2 only.

|  | Cancer risk type | Interval cancer risk | Future cancer risk<br>(excluding interval cancers at 0 to 1 year) |  |  |  | All cancer risk |
| --- | --- | --- | --- | --- | --- | --- | --- |
| Time horizon | Evaluated (y) | 0 to 1 | 1 to 2 | 1 to 3 | 1 to 4 | 1 to 5 | 0 to 5 |
| Model | Trained (y) | AUC(t) (range) |  |  |  |  |  |
| **BCSC** | 0.5 to 5 yrs | 0.58<br>(0.41,0.71) | 0.62<br>(0.55,0.68) | 0.63<br>(0.58,0.68) | 0.62<br>(0.57,0.66) | 0.60<br>(0.56,0.65) | 0.60<br>(0.56,0.65) |
| **Mirai (MIT)** | 0 to 5 yrs | 0.67<br>(0.52,0.79) | 0.61<br>(0.54,0.67) | 0.64<br>(0.58,0.68) | 0.65<br>(0.6,0.69) | 0.66<br>(0.61,0.69) | 0.66<br>(0.62,0.7) |
| **Vendor A** | 0 to 2 yrs | 0.75<br>(0.61,0.84) | 0.63<br>(0.56,0.69) | 0.68<br>(0.63,0.72) | 0.66<br>(0.62,0.7) | 0.65<br>(0.6,0.69) | 0.66<br>(0.61,0.7) |
| **Vendor B** | 0 to 1 yrs | 0.74<br>(0.57,0.83) | 0.67<br>(0.6,0.73) | 0.63<br>(0.58,0.68) | 0.6<br>(0.56,0.64) | 0.61<br>(0.57,0.65) | 0.62<br>(0.58,0.66) |
| **Vendor C** | 0 to 2 yrs | 0.66<br>(0.51,0.76) | 0.63<br>(0.55,0.69) | 0.66<br>(0.6,0.71) | 0.65<br>(0.6,0.69) | 0.65<br>(0.61,0.69) | 0.65<br>(0.61,0.69) |
| **GMIC (NYU)** | 0 to 0.25 yrs | 0.64<br>(0.48,0.75) | 0.56<br>(0.48,0.62) | 0.62<br>(0.57,0.67) | 0.53<br>(0.49,0.57) | 0.54<br>(0.49,0.58) | 0.54<br>(0.5,0.59) |

Abbreviations: AI: Artificial Intelligence; AUC(t): time-varying area under the curve; BCSC: Breast Cancer Surveillance Consortium; CAD: Computer-aided detection; GMIC: Globally-Aware Multiple Instance Classifier

## Appendix 1: Description of Artificial Algorithm

Survey questions were submitted to algorithm developers to further describe the underlying training, data, and architecture.

### Globally-Aware Multiple Instance Classifier (GMIC, NYU)

**1. Briefly describe the demographic and platform-specific composition of the data used for training: Total training sample size? Does the data contain non-white patients? Are the mammograms from multiple imaging platforms?**

*We used NYU Breast Cancer Screening Dataset to develop our model. This dataset contains 229,426 exams (1,001,093 images) from 141,472 patients who imaged at NYU Langone Health between 2010 and 2017. The dataset contains non-white patients. Mammograms were obtained on Hologic and Siemens equipment. You can find more information in this tech report*.

**2. Briefly describe the types or combination of the types of deep learning algorithms used by your model: e.g**., **CNN, DNN, GAN, etc.? Does your model utilize pretrained models or transfer learning (e.g**., **resnet, inception, etc.)?**

*The primary methodologies applied in this paper: CNN, weakly supervised learning. We used ResNet pretraining weights from ImageNet*.

**3. Briefly describe the core technologies and/or framework(s) used by your model (e.g**., **Python, Java/JVM, TensorFlow, Pytorch, Caffe, etc.)**

*PyTorch*

**4. Can your model process mammography studies beyond standard 4 views (e.g**., **unilateral only, more than 4 views, implant)?**

*Yes. It doesn’t make any assumption on the view*.

**5. How were the positive and negative labels of training images defined (e.g**., **specify the time interval from image to diagnosis, pathologically confirmed invasive cancer or DCIS, benign lesions included among negatives or as a third outcome, no known breast cancer diagnosis within 5 years, one or two subsequent negative screening exams)?**

*A breast was defined as cancer-positive if there was at least one pathology report confirming the presence of malignant lesion within 120 days of the time when the mammography images were acquired. DCIS was considered as malignant. Benign findings such as cyst and fibroadenoma were treated as another class. Cancer negative exams include exams with benign/normal findings or exams that weren’t escalated for biopsy. Please find more information in the tech report (referenced below) of this dataset*.

**6. Briefly describe your model’s input requirement (e.g**., **DICOM, PNG, “for presentation” or “for processing” view). Is any preprocessing required?**

*The implementation published on GitHub takes in a 2D / 3D matrix representation of a mammography image. The user needs to extract the image data from DICOM. Image pre-processing is included as part of the git repo*.

**7. Briefly describe your model’s output (e.g**., **does it represent the probability of cancer or can it be converted to a probability?). Does the output include anything else, such as bounding boxes or lesion segmentation?**

*The model outputs two probability scores on the presence of any benign and malignant lesion in a mammography image. The model also returns saliency maps which highlight the areas on the images that could correspond to a benign/malignant lesion. See Figure 7 of our paper as an example*.

**8. How does your model generate breast-level or patient-level predictions?**

*Breast-level predictions were calculated as the simple average over all image-level predictions*.

**9. Does your model employ any inference-time techniques, such as model ensemble or data augmentation?**

*We use model ensembling and test time data augmentation. See more details in Section 3.3.1 of our paper*.

**10. Is your model able to consider any prior exams?**

*No*

**11. Relevant citations?**

1. *The paper: https://www.sciencedirect.com/science/article/pii/S1361841520302723*
2. *NYU Breast Cancer Screening Dataset: https://cs.nyu.edu/~kgeras/reports/datav1.0.pdf*
3. *The original version of our model: https://link.springer.com/chapter/10.1007/978-3-030-32692-0_3*

**Mirai (MIT):**

**1. Briefly describe the demographic and platform-specific composition of the data used for training: Total training sample size? Does the data contain non-white patients? Are the mammograms from multiple imaging platforms?**

*The full demographics of our training set are in table 3 of the paper*.

**2. Briefly describe the types or combination of the types of deep learning algorithms used by your model: e.g**., **CNN, DNN, GAN, etc.? Does your model utilize pretrained models or transfer learning (e.g**., **resnet, inception, etc.)?**

*This is detailed in our paper*.

*Mirai leverages a ResNet to encode individual views, and a Transformer to combine multiple view representations into a patient level representation. The model was trained to predict multiple time- points simultaneously using our Additive Hazard layer, to predict traditional clinical risk factors (e.g*., *age) from the image. To make our model consistent across different mammography machines in our dataset, we used conditional adversarial training*.

**3. Briefly describe the core technologies and/or framework(s) used by your model (e.g**., **Python, Java/JVM, TensorFlow, Pytorch, Caffe, etc.)**

*Mirai was built in PyTorch and Python*

**4. Can your model process mammography studies beyond standard 4 views (e.g**., **unilateral only, more than 4 views, implant)?**

*Our model requires all four standard views (R CC, R MLO, L CC, L MLO)*

**5. How were the positive and negative labels of training images defined (e.g**., **specify the time interval from image to diagnosis, pathologically confirmed invasive cancer or DCIS, benign lesions included among negatives or as a third outcome, no known breast cancer diagnosis within 5 years, one or two subsequent negative screening exams)?**

*We trained our model to predict cancer across multiple timepoints. A patient was considered “positive” for cancer within three years if they had a pathologically confirmed invasive cancer or DCIS diagnosis within three years of their mammogram. A patient was considered “negative” for cancer within three years if they had at least three years of screening follow-up without such a diagnosis. We didn’t exclude benign lesions*.

**6. Briefly describe your model’s input requirement (e.g**., **DICOM, PNG, “for presentation” or “for processing” view). Is any preprocessing required?**

*Our code assumes For Presentation dicoms, and will convert them to pngs using the DCMTK library*

**7. Briefly describe your model’s output (e.g**., **does it represent the probability of cancer or can it be converted to a probability?). Does the output include anything else, such as bounding boxes or lesion segmentation?**

*The model outputs the probability of a cancer diagnosis within one to five years. This is represented as a 5 dimensional probability array*.

**8. How does your model generate breast-level or patient-level predictions?**

*The model generates patient-level predictions*.

**9. Does your model employ any inference-time techniques, such as model ensemble or data augmentation?**

*We do not apply model ensembling or test-time data augmentation*.

**10. Is your model able to consider any prior exams?**

*The model does not leverage prior mammograms in its predictions*.

**11. Relevant citations?**

Yala A, Mikhael PG, Strand F, et al. Toward robust mammography-based models for breast cancer risk. *Sci Transl Med*. 2021;13(578):eaba4373. doi:10.1126/scitranslmed.aba4373

#### Vendor A

**1. Briefly describe the demographic and platform-specific composition of the data used for training: Total training sample size? Does the data contain non-white patients? Are the mammograms from multiple imaging platforms?**

*We collected and curated a dataset of 1.3M images originating from Europe (France, UK) and the USA. It covers a wide range of imaging platforms, the most prominent being: Hologic, GE, Fuji, Giotto, Siemens, Philips. Images are a mixture of FFDM, DBT and 2DSM (synthetic mammography). We know from the collected centers that non-white patients are included without being able to determine precisely how many*.

**2. Briefly describe the types or combination of the types of deep learning algorithms used by your model: e.g**., **CNN, DNN, GAN, etc.? Does your model utilize pretrained models or transfer learning (e.g**., **resnet, inception, etc.)?**

*We use a mixture of 5 families of convolutional neural networks (CNN), each having a specific purpose. A first type of CNN takes a whole mammographic view as input and outputs its likelihood of malignancy. A second type of CNN extends the first one by leveraging the (lack of) symmetry between a view and its symmetrical counterpart. A third CNN is specialized in detecting all anomalies in a view, regardless of their likelihood of malignancy. A fourth CNN, further extended by a final CNN leveraging the (lack of) symmetry, takes as input high-resolution patches around detections obtained by the previous CNN and characterizes their level of suspicion. The final output of the algorithm consists in a set of positions (coordinates) within each breast view with their consolidated likelihood of malignancy obtained by fusing the image-wise and patch-wise predictions. Each model family comes with 10 instances trained by cross-validation, making a total of 50 CNNs executed on each view of each mammogram. Findings in cranial and lateral views of the same laterality are finally paired using iconic and geometrical heuristics for a more consistent output*.

**3. Briefly describe the core technologies and/or framework(s) used by your model (e.g**., **Python, Java/JVM, TensorFlow, Pytorch, Caffe, etc.)**

*The primary programming language is Python. The deep learning framework used is TensorFlow version 2. Additional machine learning methods from scikit-learn and XGBoost are also used*.

**4. Can your model process mammography studies beyond standard 4 views (e.g**., **unilateral only, more than 4 views, implant)?**

*We currently support a maximum of 4 views per mammogram, but we do support less (though off-label). Unilateral mammograms (e.g*., *L-CC and L-MLO) are supported (symmetric models are disabled in this case). Unique views (e.g*., *L-CC and R-CC) are also supported. Additional screening views such as ML, LM, XCC are also supported. In case of duplicated views (e.g*., *two L-CC), the most recent image is selected (we hypothesize that in case of duplicated views the most recent is a reshoot due to bad quality of the older version)*.

**5. How were the positive and negative labels of training images defined (e.g**., **specify the time interval from image to diagnosis, pathologically confirmed invasive cancer or DCIS, benign lesions included among negatives or as a third outcome, no known breast cancer diagnosis within 5 years, one or two subsequent negative screening exams)?**

*Positive cases were confirmed by a positive biopsy (either invasive cancer or DCIS) within 24 months from screening date. For each mammogram, the cancer presence was confirmed (annotated) by an expert radiologist to avoid injecting interval cancers in the training set. Regarding negative cases, those were confirmed by a negative or benign screening exam within 24 months after the screening date. Cases with benign lesions (as confirmed after a diagnostic mammogram or a biopsy) were included in the same group as negative cases. Cases with known history of breast cancer (regardless of when it happened) and breast surgery were excluded*.

**6. Briefly describe your model’s input requirement (e.g**., **DICOM, PNG, “for presentation” or “for processing” view). Is any preprocessing required?**

*We use the FOR PRESENTATION DICOM images for the AI analysis. No additional preprocessing is required. Images are normalized internally (i.e*., *by the algorithm) to make them look similar across vendors. The normalization procedure is not disclosed*.

**7. Briefly describe your model’s output (e.g**., **does it represent the probability of cancer or can it be converted to a probability?). Does the output include anything else, such as bounding boxes or lesion segmentation?**

*[Vendor A algorithm] outputs 2 scores: a raw score, ranging between 0 and 1, and a discretized score on a 1-10 scale to ease its interpretation. The score was calibrated on a screening distribution with a cancer prevalence of 5:1000 so it can directly be interpreted as a probability. Additionally, [Vendor A algorithm] returns the detection bounding boxes. No pixelic lesion segmentation is performed*.

**8. How does your model generate breast-level or patient-level predictions?**

*The breast-level score is obtained as the highest score of the detected lesions in the breast. The patient-level score is the highest score of the left and right breast. Therefore, it is always possible to connect the score at any level to a specific lesion in a specific view, which eases interpretability*.

**9. Does your model employ any inference-time techniques, such as model ensemble or data augmentation?**

*Given the multiplicity of our CNN, we extensively use ensembling techniques (bagging). Each model family has 10 instances, which are combined (averaged) to form a unique prediction for that family. No test-time augmentation is done*.

**10. Is your model able to consider any prior exams?**

*The use of prior examinations is currently under development. We have very promising preliminary results on certain model families and are currently extending it to other families*.

#### Vendor B

**1. Briefly describe the demographic and platform-specific composition of the data used for training: Total training sample size? Does the data contain non-white patients? Are the mammograms from multiple imaging platforms?**

*In construction of the model, we utilized data from four primary vendors: Hologic, GE, Giotto, and Siemens. The model has been trained on >4M images. The dataset contains Caucasian, Asian, Hispanic, and African American demographics*.

**2. Briefly describe the types or combination of the types of deep learning algorithms used by your model: e.g**., **CNN, DNN, GAN, etc.? Does your model utilize pretrained models or transfer learning (e.g**., **resnet, inception, etc.)?**

*Our models include convolutional neural networks comprising several customized architectures. We also construct an ensemble of several of these models with various training parameters to construct the final model. We use transfer learning on internal data to make our optimization process more efficient. We don’t use any publicly pretrained models to avoid potential bias*.

**3. Briefly describe the core technologies and/or framework(s) used by your model (e.g**., **Python, Java/JVM, TensorFlow, Pytorch, Caffe, etc.)**

*The product is built as a JVM service-based tool using dcmtk to directly interface to DICOM compatible devices. The model is built using TensorFlow as the primary framework for deep learning, with several proprietary customizations to improve the performance of our models*.

**4. Can your model process mammography studies beyond standard 4 views (e.g**., **unilateral only, more than 4 views, implant)?**

*The algorithm assesses 4 standard view images [CC-L, CC-R, MLO-L, MLO-R]. When the 4 standard views are a subset of more than 4 images in a case, the algorithm utilizes a custom image selection heuristic to determine an overall result based on the appropriate 4-image subset. The heuristic can be customized to fit local processes. We believe it is critical that our deployments are executed with understanding of how each local site works (e.g*., *how the images are produced). This information is then used to ensure the algorithm is optimally deployed. Versions of this heuristic have already been used successfully in independent analyses*.

**5. How were the positive and negative labels of training images defined (e.g**., **specify the time interval from image to diagnosis, pathologically confirmed invasive cancer or DCIS, benign lesions included among negatives or as a third outcome, no known breast cancer diagnosis within 5 years, one or two subsequent negative screening exams)?**

*Our positive definition is based on pathology or surgical follow up within 6 months or 12 months as proof of malignancy. Our negative definition has a requirement of negative at screening plus 24 to 36 months follow up with a negative result*.

**6. Briefly describe your model’s input requirement (e.g**., **DICOM, PNG, “for presentation” or “for processing” view). Is any preprocessing required?**

*We only report on diagnosis-grade “For Presentation” Images as described in the DICOM standard. No preprocessing is required*.

**7. Briefly describe your model’s output (e.g**., **does it represent the probability of cancer or can it be converted to a probability?). Does the output include anything else, such as bounding boxes or lesion segmentation?**

*We provide a binary decision of Recall or No Recall based on a calibration step with each provider’s local data environment. Included in a recall decision would be side-wise (L/R) and view-wise (MLO/CC) details. We also provide an explanatory ROI for informing the logic behind the recall decision. We can provide a score; however, it is our belief that the score is not reflective of the probability of cancer*.

**8. How does your model generate breast-level or patient-level predictions?**

*We generate a prediction at the image level across an ensemble of models and then apply a reduction heuristic to achieve the final result. The recall decision includes a case-wise, side-wise and view-wise output*.

**9. Does your model employ any inference-time techniques, such as model ensemble or data augmentation?**

*We use a varied ensemble of ML models. We found that, with our models, inference-time techniques such as test time augmentation were not needed to achieve very good performance. This allows us to be time and cost efficient at inference time*.

**10. Is your model able to consider any prior exams?**

*Some of our internal models have been trained using priors and we have seen strong performance improvements. The production model doesn’t not consider any prior exams at inference-time yet, but this feature is actively being worked on and is expected to be released in 2022*.

#### Vendor D

**1. Briefly describe the demographic and platform-specific composition of the data used for training: Total training sample size? Does the data contain non-white patients? Are the mammograms from multiple imaging platforms?**

*Predominantly Caucasian women were included in the training. Approximately one thousand breast cancer cases and ten thousand controls were used from multiple mammography machine vendors*.

**2. Briefly describe the types or combination of the types of deep learning algorithms used by your model: e.g**., **CNN, DNN, GAN, etc.? Does your model utilize pretrained models or transfer learning (e.g**., **resnet, inception, etc.)?**

*The deep learning algorithm is based on a combination of inception-based convolutional neural networks and U-Net. Transfer learning techniques and regularization were also used during model training*.

**3. Briefly describe the core technologies and/or framework(s) used by your model (e.g**., **Python, Java/JVM, TensorFlow, Pytorch, Caffe, etc.)**

*Caffe and Tensorflow are used for model training and deployment*.

**4. Can your model process mammography studies beyond standard 4 views (e.g**., **unilateral only, more than 4 views, implant)?**

*The model processes 4 standard views with and without implants*.

**5. How were the positive and negative labels of training images defined (e.g**., **specify the time interval from image to diagnosis, pathologically confirmed invasive cancer or DCIS, benign lesions included among negatives or as a third outcome, no known breast cancer diagnosis within 5 years, one or two subsequent negative screening exams)?**

*The time from mammogram to diagnosis or end of follow-up was up to 4 years based on pathology confirmed invasive and in-situ cancers. Benign lesions were included for controls. The proportions of invasive, in-situ, benign lesions, normals matched a European screening population*.

**6. Briefly describe your model’s input requirement (e.g**., **DICOM, PNG, “for presentation” or “for processing” view). Is any preprocessing required?**

*The model requires for presentation DICOM images and age from the image tags*.

**7. Briefly describe your model’s output (e.g**., **does it represent the probability of cancer or can it be converted to a probability?). Does the output include anything else, such as bounding boxes or lesion segmentation?**

*The model outputs absolute risk of breast cancer in the population adjusted for age of the woman. The model also outputs the average risk of women at the same age*.

**8. How does your model generate breast-level or patient-level predictions?**

*Artificial intelligence analyses four images and image feature relationships between the four images. The model further uses breast cancer incidence rates and competing risks from the general population when predicting population based absolute risk of breast cancer for the woman*.

**9. Does your model employ any inference-time techniques, such as model ensemble or data augmentation?**

*The model uses ensemble techniques, and the model uses a time-to-event model with adjustment for competing risks*.

**10. Is your model able to consider any prior exams?**

*The model is designed for analyzing images prior to breast cancer. It analyzes images at one time point*.

